# Association between regular antithrombotic drugs use and Nonbiliary Acute Pancreatitis risk: a prospective cohort study

**DOI:** 10.64898/2025.12.22.25342783

**Authors:** Chenyang Wei, Ningning Mi, Jinyu Zhao, Pengfei Li, Zhengping An, Sheng Chen, Yanyan Lin, Ping Yue, Jinqiu Yuan, Wenbo Meng

**Author notes:** These authors are the corresponding authors (Jinqiu Yuan, Wenbo Meng). Chenyang Wei and Ningning Mi have contributed equally to this work. **Correspondence:** Wenbo Meng, Department of General Surgery, The First Hospital of Lanzhou University, Lanzhou, 730000, Gansu, China.; Jinqiu Yuan, Clinical Research Center, Big Data Center, The Seventh Affiliated Hospital, Sun Yat-sen University, Shenzhen, 518107, Guangdong, China.

## Abstract

**BACKGROUND:** Antithrombotic drugs are widely used for the prevention of cardiovascular disease; however, their association with nonbiliary acute pancreatitis remains unclear. To investigate the association between regular use of antithrombotic drugs and the risk of developing nonbiliary acute pancreatitis.

**METHODS:** This prospective cohort study included 431,754 participants who did not have acute pancreatitis, gallstones, or cancer. Incident nonbiliary acute pancreatitis cases were identified through primary healthcare and hospitalization data, excluding gallstone-related cases. Cox proportional hazards models, adjusted for sociodemographic characteristics, lifestyle, comorbidities, and medication, were used to estimate the association with regular antithrombotic drug use.

**RESULTS:** During a median follow-up period of 13.73 years, 2,189 cases of nonbiliary acute pancreatitis were recorded. Users of antithrombotic drugs had a 31% higher risk of developing nonbiliary acute pancreatitis than non-users (hazard ratio [HR], 1.31; 95% confidence interval [CI], 1.08-1.61; p=0.007). Risk varied by specific agent: clopidogrel was associated with a 53% increased risk (HR, 1.53; 95% CI, 1.08-2.16; p=0.016). For warfarin, the overall association was not statistically significant (HR, 1.32; 95% CI, 0.97-1.80; p=0.076); however, subgroup analysis indicated that the association was confined to participants without diabetes (P for interaction, 0.015; HR, 1.64; 95% CI, 1.15-2.35; p=0.007). There was no significant association between low-dose aspirin and use of dipyridamole.

**CONCLUSION:** Regular use of antithrombotic drugs, particularly clopidogrel, was associated with an increased risk of developing nonbiliary acute pancreatitis. The risk associated with warfarin was specific to participants without diabetes.

## INTRODUCTION

Acute pancreatitis (AP) is an inflammatory disorder of the pancreas, which is associated with substantial morbidity and mortality, and it imposes a substantial burden on healthcare systems worldwide [1]. In severe cases, it can progress to systemic inflammation and multiple organ failure, with a mortality rate as high as 85% [2]. The etiological spectrum of AP has significant geographical variation; in Western countries, biliary disease is the leading cause, followed by alcoholic and hypertriglyceridemia-induced pancreatitis. However, in Asian countries, including China, hypertriglyceridemia accounts for a substantially higher percentage of cases [3]. The rapidly increasing prevalence of hypertriglyceridemia-related AP has established it as a pressing global health concern [4,5].

In addition to the traditional causes, medications are recognized as potentially inducing AP [6,7]. Antithrombotic agents have drawn particular attention due to their widespread use in patients with cardiovascular diseases [8,9]. However, evidence of the association between the use of these drugs and the risk of developing AP remains controversial, and consensus has not yet been reached. Evidence from animal models suggests that there is a potential protective effect; pre-treatment with low-dose warfarin can alleviate ischemia/reperfusion-induced pancreatic injury via this mechanism [10,11]. A recent large-scale clinical study also reported that among patients with established AP, the use of low-dose aspirin was associated with lower mortality and a reduced risk of complications [12]. In contrast, a study had reported that actively using clopidogrel were associated with an increased risk of acute pancreatitis [13]. This conflicting body of evidence creates a dilemma for clinicians who are managing a substantial cohort of patients with cardiovascular disease, and who require long-term antithrombotic therapy, making it challenging to assess their potential risk of developing AP.

Given these conflicting findings, a rigorous investigation is warranted to clarify the relationship between the use of antithrombotic drugs and the risk of developing AP. If these drugs be confirmed as a risk factor, their cardiovascular benefits must be rigorously weighed against the risk of developing AP, in clinical decision-making, necessitating increased clinical vigilance among prescribers. Conversely, evidence of a protective effect would justify further studies into the mechanisms of action. To address this knowledge gap, a prospective analysis of the association between the use of antithrombotic drugs and the risk of developing AP, in a large cohort of 431,754 individuals, was conducted.

## MATERIALS AND METHODS

### Study cohort

Data from the study cohort was obtained from the United Kingdom (UK) Biobank, a large prospective cohort of >500,000 participants, aged 37-73 years. The cohort was established between 2006 and 2010 across 22 assessment centers in England, Wales, and Scotland. The participants were followed-up regularly for data updates. During recruitment, all participants completed a baseline questionnaire and underwent assessments that included physical measurements and detailed information on lifestyle and use of medication. Further details regarding the UK Biobank cohort have been published elsewhere [14]. Participants with a baseline diagnosis of cancer, gallstone disease, or acute pancreatitis were excluded. To prevent gallstone disease from confounding the results, participants who developed acute AP during follow-up, but had previously been diagnosed with gallstone disease, were excluded. Participants with incomplete follow-up data were also excluded. The final analytical cohort comprised 431,754 participants (Supplementary Figure 1). All diagnoses were based on the International Classification of Diseases-10 (ICD-10) diagnostic criteria.

### Assessment of exposure

The primary exposure was defined as regular use of antithrombotic drugs, at baseline, including anticoagulants and antiplatelet drugs. The specific medications that were considered were: warfarin, low-dose aspirin, clopidogrel, and dipyridamole. At baseline, the regular use of antithrombotic drugs was first assessed from participants using a touchscreen questionnaire, and confirmed during verbal interviews with trained staff. In the touchscreen questionnaire, participants were asked, “Do you regularly take any other prescription medications?”. “Regular use” of prescription medications was defined as taking them on a long-term and scheduled basis, whether daily, weekly, or monthly (e.g., depot injections), as reported during the verbal interview. Short-term medications, such as courses of antibiotics, were excluded. If the participant selected “Yes” or “Do not know”, the interviewer would ask; “In the touchscreen, you said you are taking regular prescription medications. Can you now tell me what these are?”. Information regarding the use of antithrombotic drugs was recorded using a coded list. The antithrombotic drugs recorded included warfarin, low-molecular-weight heparin, low-dose aspirin, clopidogrel, and dipyridamole. Information regarding the dose and duration of use of antithrombotic agents was not collected. Detailed questions on the use of antithrombotic agents can be found on the UK Biobank website.

### Ascertainment of outcome

Incident cases of AP were identified from hospital records linked to the Health and Social Care Information Centre (England and Wales) and the National Health Service Central Register (Scotland), using the ICD-10 code, K85. The diagnosis of AP was further validated if at least two of the following three criteria were fulfilled: (1) characteristic abdominal pain, (2) serum amylase and/or lipase levels greater than three times the upper limit of normal, and (3) imaging findings consistent with AP on contrast-enhanced computed tomography or magnetic resonance imaging. Among the participants with AP, nonbiliary AP was confirmed by excluding those with a concomitant or prior diagnosis of gallstone disease. Assessment of the severity of disease was not performed due to the lack of detailed clinical and laboratory data in the databases.

### Assessment of covariates

At enrollment, information on covariates was collected through the touchscreen questionnaire and face-to-face interviews, covering socio-demographic characteristics, anthropometric measurements, lifestyle factors, medical history, and use of medication. Specifically, the data included: (1) socio-demographic factors: age, sex, ethnicity and socioeconomic status (measured using the Index of Multiple Deprivation [IMD], directly obtained from the UK Biobank); (2) anthropometric data: height and weight, and body mass index (BMI), calculated as kg/m²; (3) lifestyle factors: smoking status, alcohol consumption, physical activity (assessed using the short-form International Physical Activity Questionnaire, and healthy diet (defined according to American Heart Association guidelines; Supplementary Table 2); (4) medical history: type 2 diabetes, hypertension, hyperlipidemia, and cardiovascular disease; (5) use of medication: nonsteroidal antiinflammatory drugs (NSAIDs), hypolipidemic drugs, multivitamin and mineral supplements.

### Statistical analysis

Baseline characteristics are presented as mean (standard deviation, SD) for continuous variables and as count (percentage) for categorical variables. Person-years of follow-up were calculated from the date of assessment for each participant until the first diagnosis of AP, death, or the end of follow-up (May 31, 2022), whichever was first.

Cumulative incidence curves for nonbiliary AP were generated to compare non-users of antithrombotic drugs with users of antithrombotic drugs, including anticoagulants, antiplatelet agents, and specific medications. Initially, a crude model was constructed to determine the association between the use of antithrombotic drugs and the risk of developing nonbiliary AP. Next, three adjusted models were generated: Model 1 was adjusted for age, sex, ethnicity, and IMD; Model 2 was further adjusted for BMI, smoking, alcohol consumption, physical activity, and healthy diet; and Model 3 was additionally adjusted for the presence of type 2 diabetes, hypertension, or hyperlipidemia, use of NSAIDs, use of hypolipidemic drugs, and use of multivitamins and mineral supplements. The proportional hazards assumption was verified using Schoenfeld residuals for all models, and there were no violations.

Subgroup analyses were performed according to pharmacological class (anticoagulants or antiplatelet agents) and individual drugs (warfarin, aspirin, clopidogrel, and dipyridamole). The results from the most fully adjusted model (Model 3) are reported as the primary findings of this study.

The potential moderating effects of age, sex, BMI, smoking status, alcohol consumption, physical activity, healthy diet, presence of type 2 diabetes, hypertension, or hyperlipidemia, NSAID intake, use of hypolipidemic drugs, and use of multivitamins and mineral supplements, were evaluated using stratified analysis and interaction testing with a likelihood ratio test. Given that the presence of type 2 diabetes and hyperlipidemia suggested that there was a potential effect modification in the overall analysis, their interaction effects were tested in the aforementioned stratified medication analyses.

To evaluate the robustness of the findings, several sensitivity analyses were performed: (1) further adjustment for history of cardiovascular diseases in Model 3, as Model 4, to assess the extent to which the association could be explained by pre-existing cardiovascular conditions; (2) exclusion of participants who developed the outcome within the first year of follow-up, to minimize reverse causality; and (3) a propensity score matching analysis to evaluate the effect of the use of antithrombotic drugs on the risk of developing nonbiliary AP.

All analyses were performed using R version 4.5.1. Statistical significance was set as two-tailed p<0.05.

## RESULTS

### Baseline characteristics of participants

Table 1 presents the baseline characteristics of the study cohort, stratified by use of antithrombotic drugs. There were 431,754 participants, of whom 11,140 used antithrombotic drugs. Among these, 4,452 used anticoagulants only, 6,610 used antiplatelet agents only, and 78 used antiplatelet agents and anti-coagulants. Regarding specific medications, 4,397 used warfarin, 2,514 used low-dose aspirin, 950 used dipyridamole, 2,818 used clopidogrel, and 93 used aspirin and clopidogrel. Users of antithrombotic drugs were more likely to be older, male, smokers, consumers of alcohol, and have a higher prevalence of comorbidities, including type 2 diabetes, hypertension, and hyperlipidemia.

**Table 1.**
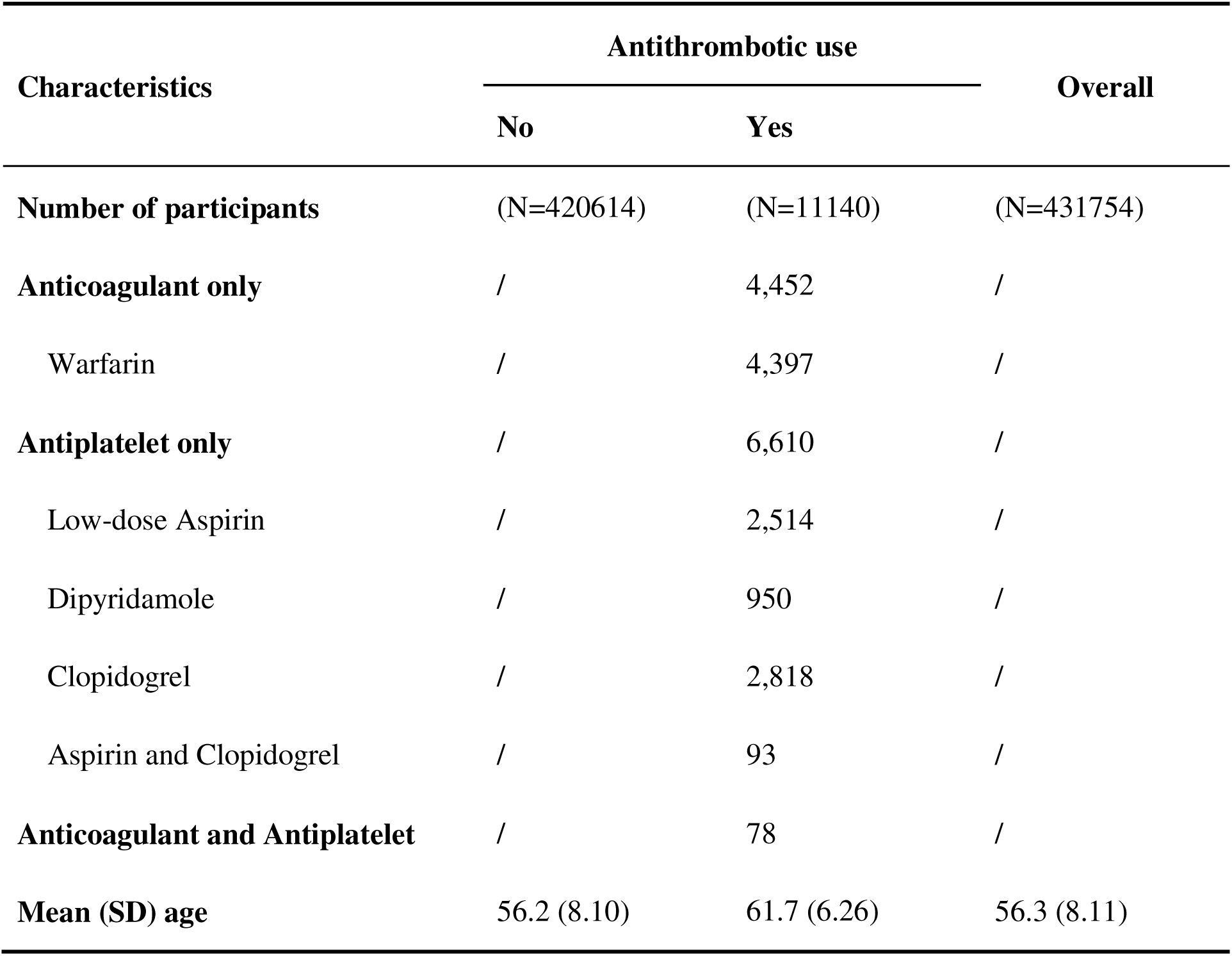
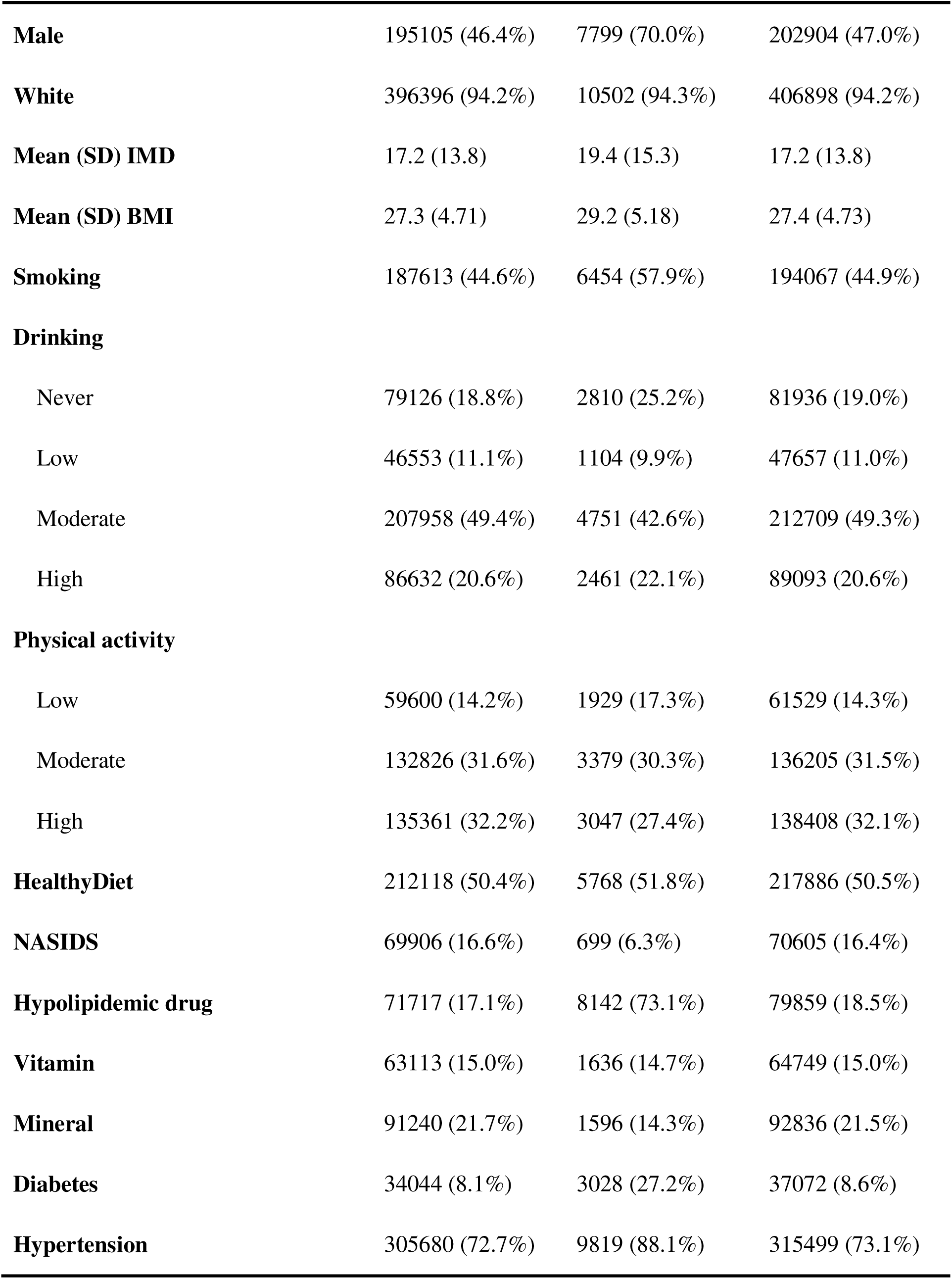
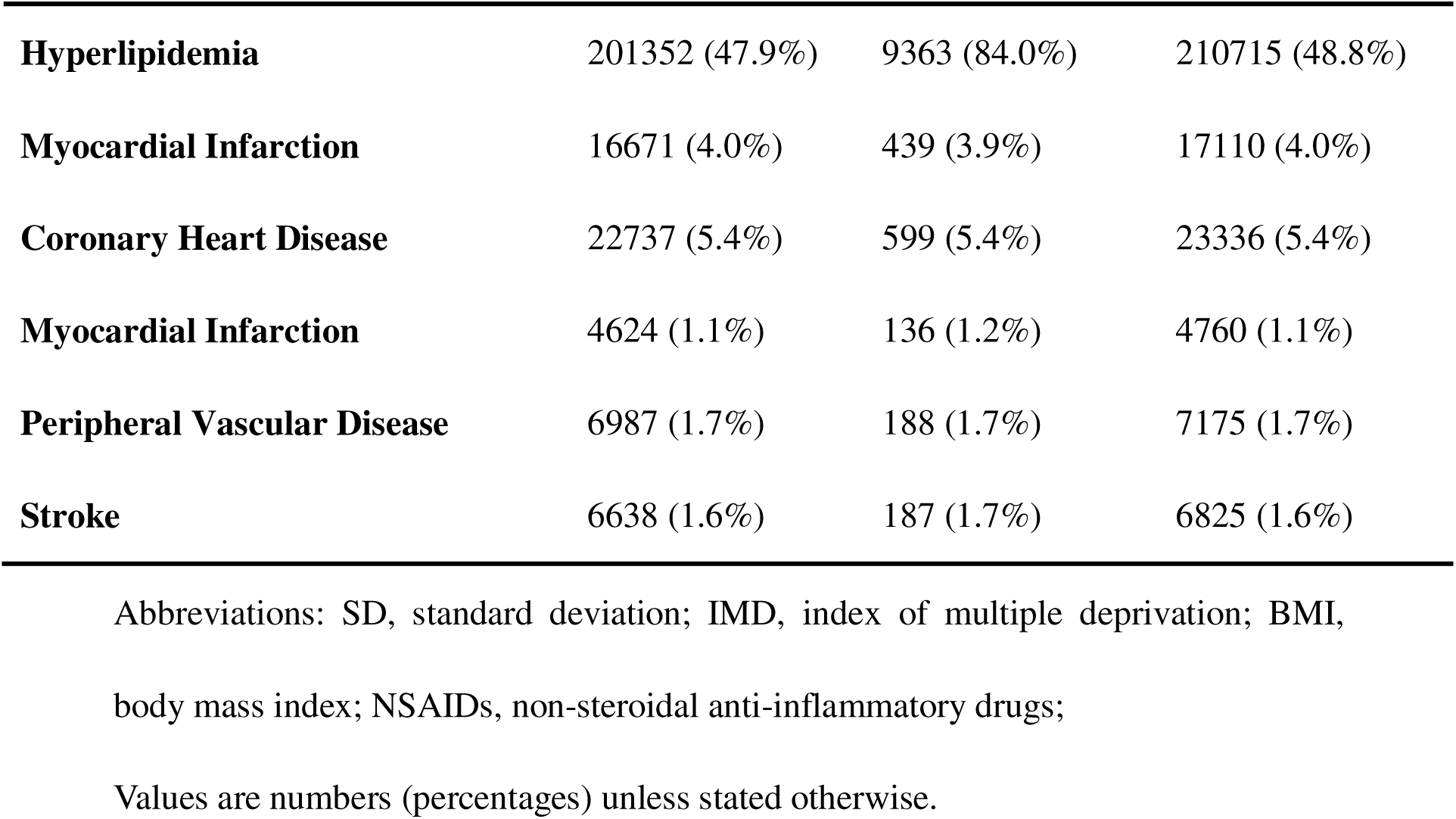
Baseline characteristics of participants by antithrombotic use.

| Characteristics | Antithrombotic use |  | Overall |
| --- | --- | --- | --- |
|  | No | Yes |  |
| **Number of participants** | (N=420614) | (N=11140) | (N=431754) |
| **Anticoagulant only** | / | 4,452 | / |
| Warfarin | / | 4,397 | / |
| **Antiplatelet only** | / | 6,610 | / |
| Low-dose Aspirin | / | 2,514 | / |
| Dipyridamole | / | 950 | / |
| Clopidogrel | / | 2,818 | / |
| Aspirin and Clopidogrel | / | 93 | / |
| **Anticoagulant and Antiplatelet** | / | 78 | / |
| **Mean (SD) age** | 56.2 (8.10) | 61.7 (6.26) | 56.3 (8.11) |
| **Male** | 195105 (46.4%) | 7799 (70.0%) | 202904 (47.0%) |
| **White** | 396396 (94.2%) | 10502 (94.3%) | 406898 (94.2%) |
| **Mean (SD) IMD** | 17.2 (13.8) | 19.4 (15.3) | 17.2 (13.8) |
| **Mean (SD) BMI** | 27.3 (4.71) | 29.2 (5.18) | 27.4 (4.73) |
| **Smoking** | 187613 (44.6%) | 6454 (57.9%) | 194067 (44.9%) |
| **Drinking** |  |  |  |
| Never | 79126 (18.8%) | 2810 (25.2%) | 81936 (19.0%) |
| Low | 46553 (11.1%) | 1104 (9.9%) | 47657 (11.0%) |
| Moderate | 207958 (49.4%) | 4751 (42.6%) | 212709 (49.3%) |
| High | 86632 (20.6%) | 2461 (22.1%) | 89093 (20.6%) |
| **Physical activity** |  |  |  |
| Low | 59600 (14.2%) | 1929 (17.3%) | 61529 (14.3%) |
| Moderate | 132826 (31.6%) | 3379 (30.3%) | 136205 (31.5%) |
| High | 135361 (32.2%) | 3047 (27.4%) | 138408 (32.1%) |
| **HealthyDiet** | 212118 (50.4%) | 5768 (51.8%) | 217886 (50.5%) |
| **NASIDS** | 69906 (16.6%) | 699 (6.3%) | 70605 (16.4%) |
| **Hypolipidemic drug** | 71717 (17.1%) | 8142 (73.1%) | 79859 (18.5%) |
| **Vitamin** | 63113 (15.0%) | 1636 (14.7%) | 64749 (15.0%) |
| **Mineral** | 91240 (21.7%) | 1596 (14.3%) | 92836 (21.5%) |
| **Diabetes** | 34044 (8.1%) | 3028 (27.2%) | 37072 (8.6%) |
| **Hypertension** | 305680 (72.7%) | 9819 (88.1%) | 315499 (73.1%) |
| **Hyperlipidemia** | 201352 (47.9%) | 9363 (84.0%) | 210715 (48.8%) |
| **Myocardial Infarction** | 16671 (4.0%) | 439 (3.9%) | 17110 (4.0%) |
| **Coronary Heart Disease** | 22737 (5.4%) | 599 (5.4%) | 23336 (5.4%) |
| **Myocardial Infarction** | 4624 (1.1%) | 136 (1.2%) | 4760 (1.1%) |
| **Peripheral Vascular Disease** | 6987 (1.7%) | 188 (1.7%) | 7175 (1.7%) |
| **Stroke** | 6638 (1.6%) | 187 (1.7%) | 6825 (1.6%) |

Abbreviations: SD, standard deviation; IMD, index of multiple deprivation; BMI, body mass index; NSAIDs, non-steroidal anti-inflammatory drugs; Values are numbers (percentages) unless stated otherwise.

### Use of antithrombotic drugs and the risk of developing Nonbiliary AP

During a median follow-up of 13.73 years, 2,189 new participants with nonbiliary AP were recorded, including 109 who used antithrombotic drugs and 2,080 who did not use antithrombotic drugs. Compared with non-users, there was a higher cumulative incidence of AP in users, particularly among users of anticoagulants and clopidogrel (Figure 1a, b, and c).

**Figure 1.**
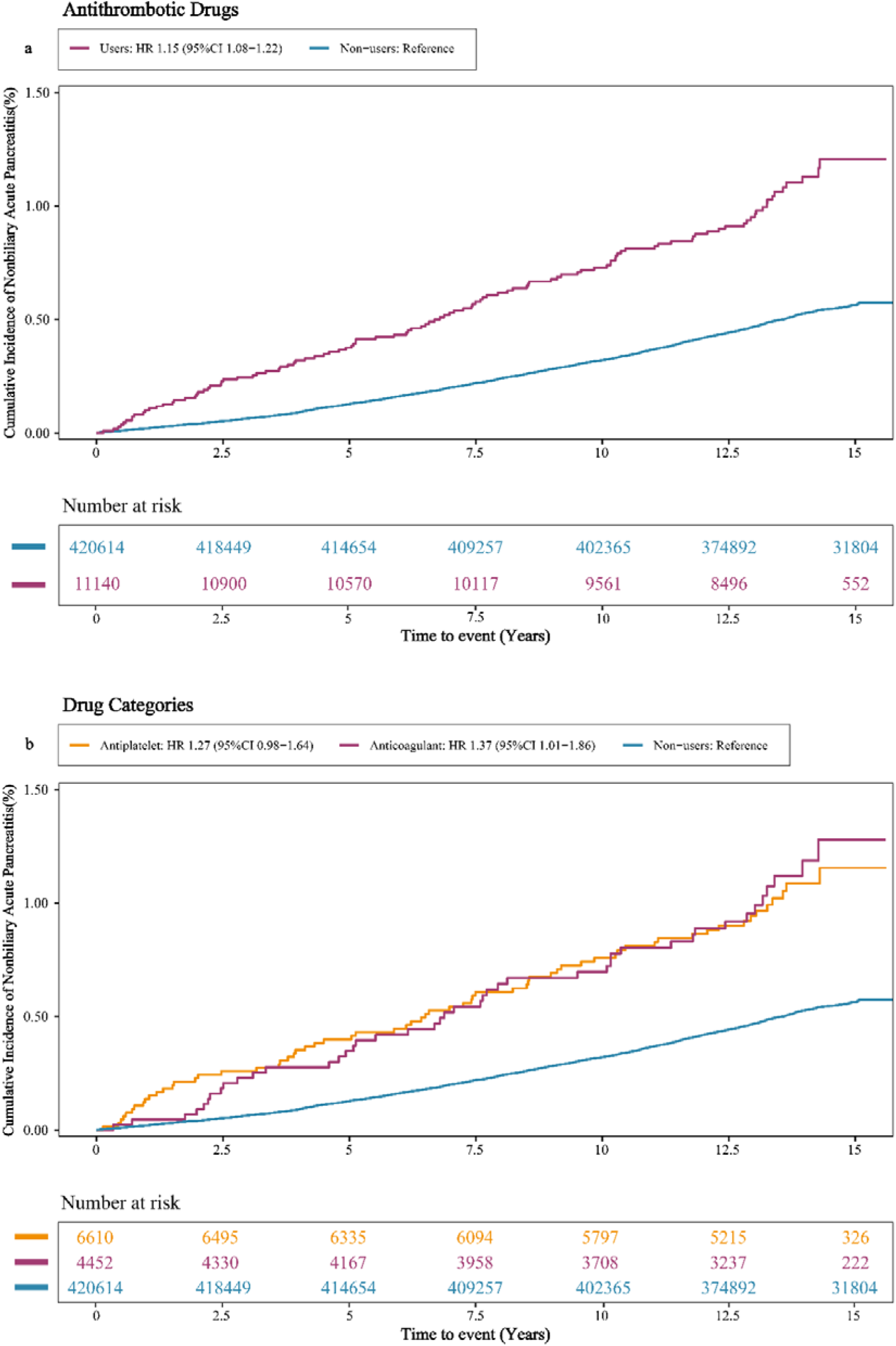
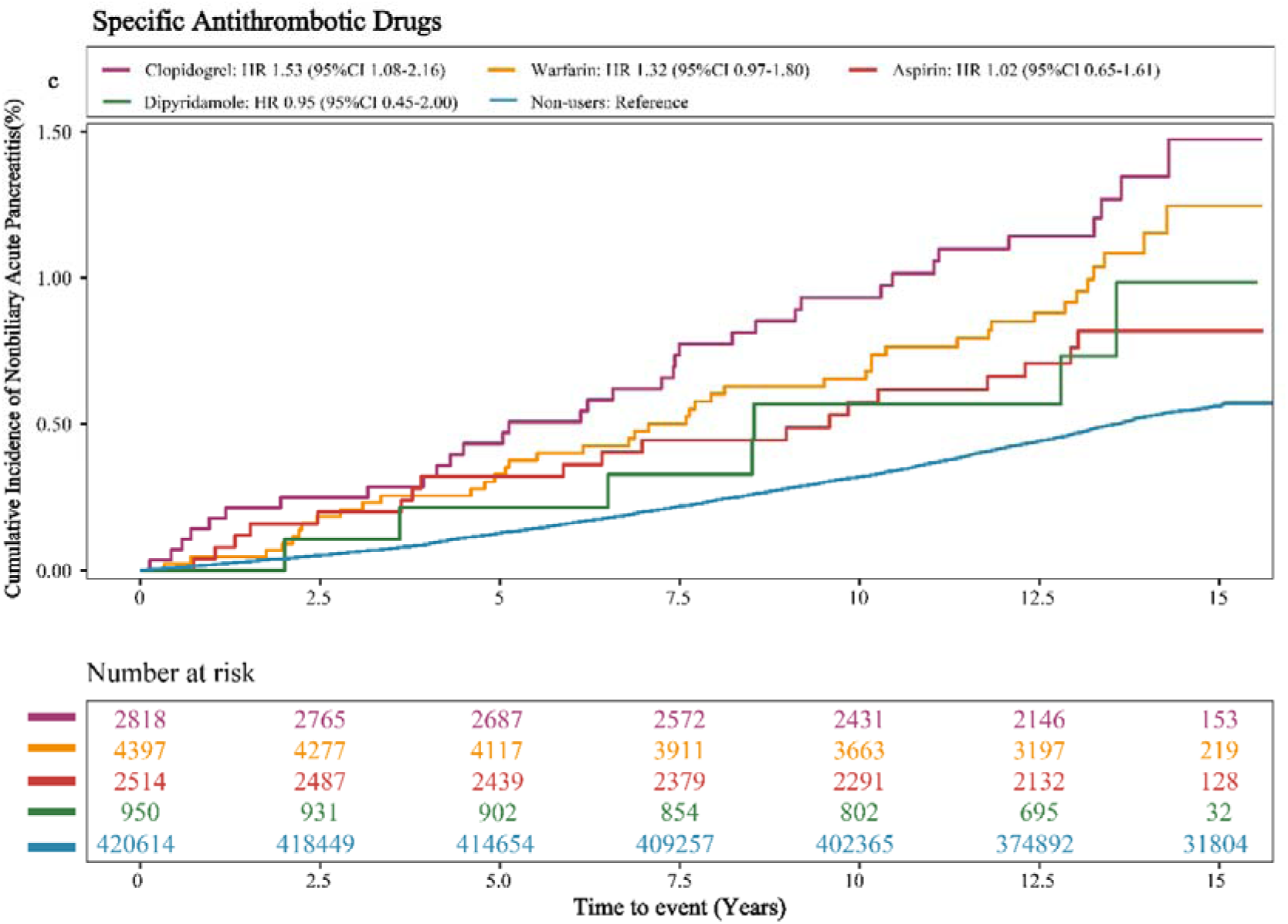
Cumulative Incidence of Nonbiliary AP Among Users of Antithrombotic Drugs with Non-Users. Estimated effects were calculated using the Cox model adjusted by age, gender, ethnicity, IMD, BMI, smoking status, alcohol intake, physical activity, dietary characteristics, type 2 diabetes, hypertension, hyperlipidemia, NSAID intake, Hypolipidemic drug intake, multivitamin use and intake of mineral supplements. a shows the cumulative incidence of nonbiliary acute pancreatitis in overall antithrombotic drug users and non-users. b shows the cumulative incidence of nonbiliary acute pancreatitis in antiplatelet drug users, anticoagulant drug users, and non-users. c shows the cumulative incidence of nonbiliary acute pancreatitis in clopidogrel, warfarin, aspirin, dipyridamole users, and non-users. **Abbreviations:** IMD, index of multiple deprivation; BMI, body mass index; NSAID, nonsteroidal anti-inflammatory drug; AP, acute pancreatitis.

In the crude model, the use of antithrombotic drugs was associated with a 74% increased risk of developing nonbiliary AP (hazard ratio [HR], 1.74; 95% confidence interval [CI], 1.44-2.12; p<0.001). After adjusting for age, sex, ethnicity, and IMD (Model 1), the HR was 1.66 (95% CI, 1.37-2.02; p<0.001). The association attenuated somewhat but remained significant after further adjustment for lifestyle factors (Model 2) (HR, 1.44; 95% CI, 1.18-1.75; p<0.001). In Model 3, which was further adjusted for comorbidities and use of medication, the result did not change substantially (HR, 1.31; 95% CI, 1.08-1.61; p=0.007) (Table 2). In the analysis by pharmacological class, use of anticoagulants had a significantly increased risk (HR, 1.37; 95% CI, 1.01-1.86; p=0.040). Specifically, warfarin showed a trend towards an increased risk, although it did not reach statistical significance (HR, 1.32; 95% CI, 0.97-1.80; p=0.076). Analysis of other anticoagulants could not be performed or yielded imprecise results due to limitations in sample size. The overall association for antiplatelet agents was at the margin of statistical significance (HR, 1.27; 95% CI, 0.98-1.64; p=0.071). The use of clopidogrel was associated with a significantly increased risk (HR, 1.53; 95% CI, 1.08-2.16; p=0.016), whereas aspirin (HR, 1.02; 95% CI, 0.65-1.61; p=0.934) and dipyridamole (HR, 0.95; 95% CI, 0.45-2.00; p=0.890) did not have a significant association (Figure 2).

**Figure 2.**
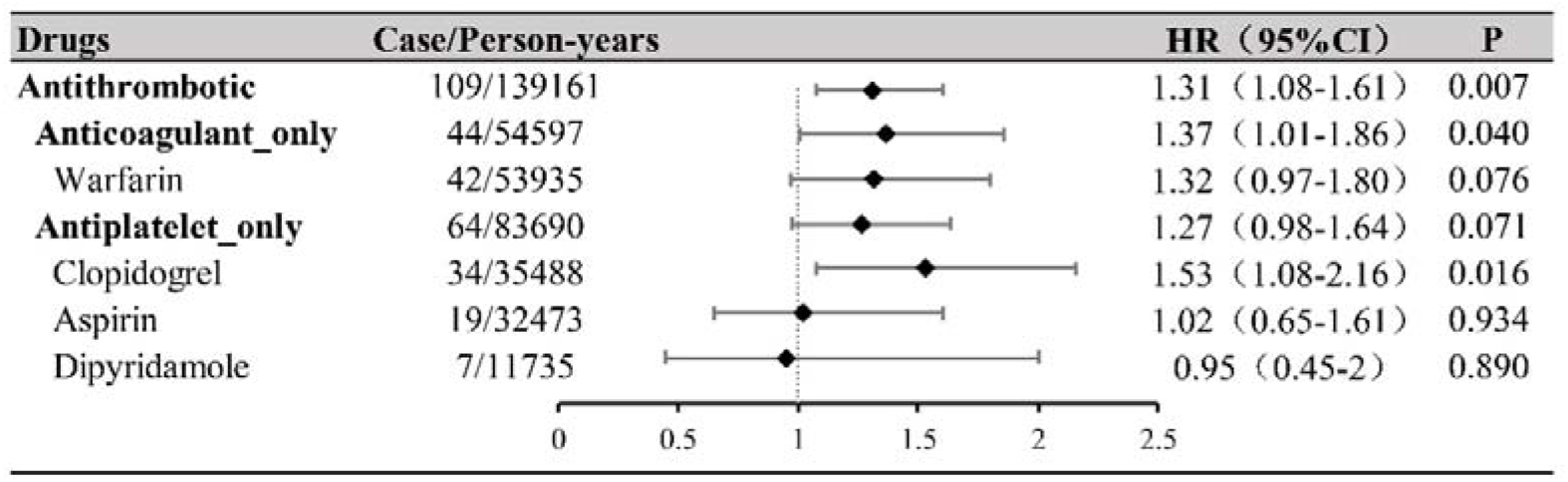
Forest plot for the association of antithrombotic drug use with the risk of Nonbiliary AP. The model was adjusted for age, gender, ethnicity, IMD, BMI, smoking status, alcohol intake, physical activity, dietary characteristics, type 2 diabetes, hypertension, hyperlipidemia, NSAID intake, hypolipidemic drug intake, multivitamin use, and mineral supplement intake; **Abbreviations:** IMD, the Index of Multiple Deprivation; BMI, body mass index; NSAID, nonsteroidal anti-inflammatory dr

**Table 2.** Associations between Antithrombotic use and risk of Nonbiliary Acute Pancreatitis(n=431,754).

| Variables | Antithrombotic use |  | p-value |
| --- | --- | --- | --- |
|  | No ( N=420,614 ) | Yes (N=11,140) |  |
| **Case/person-years** | 2,080 / 5,652,141 | 109 / 139,161 |  |
| **IR (per 10,000 person-years)** | 3.68 | 7.83 |  |
| **Multivariate-adjusted HR (95% CI)** |  |  |  |
| **Crude model** | 1.00 (Reference) | 1.74 ( 1.44-2.12 ) | <0.001 |
| **model 1** | 1.00 (Reference) | 1.66 ( 1.37-2.02 ) | <0.001 |
| **model 2** | 1.00 (Reference) | 1.44 ( 1.18-1.75 ) | <0.001 |
| **model 3** | 1.00 (Reference) | 1.31 ( 1.08-1.61 ) | 0.007 |

### Subgroup analysis

The analysis revealed significant interactions; the association between overall use of antithrombotic drugs and the risk of developing nonbiliary AP varied depending on the presence of diabetes (P for interaction = 0.001) or hyperlipidemia (P for interaction = 0.044). Specifically, among participants without diabetes, the use of antithrombotic drugs was significantly associated with an increased risk of developing nonbiliary AP (HR, 1.52; 95% CI, 1.19-1.96; p=0.001). However, this association was not observed for participants with diabetes (HR, 1.08; 95% CI, 0.78-1.50; p=0.644). The increased risk was more pronounced for participants without hyperlipidemia (HR, 1.96; 95% CI, 1.22-3.15; p=0.006), while the association was weaker and did not reach statistical significance for participants with hyperlipidemia (HR, 1.23; 95% CI, 0.99-1.53; p=0.063) (Figure 3).

**Figure 3.**
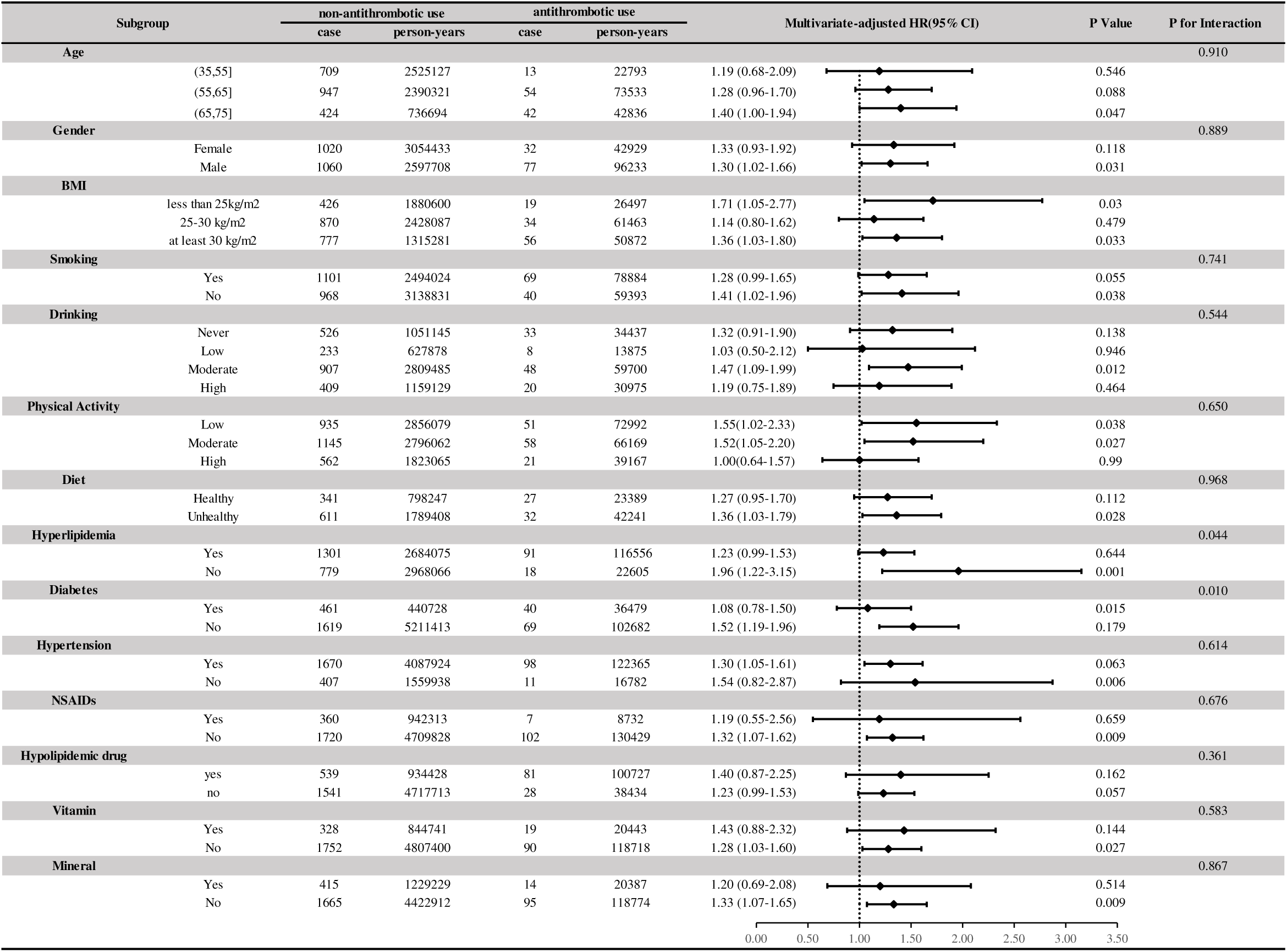
Subgroup analyses of Antithrombotic Drugs Use and the risk of Nonbiliary AP. Estimated effects were calculated using the Cox model adjusted by age, gender, ethnicity, IMD, BMI, smoking status, alcohol intake, physical activity, dietary characteristics, type 2 diabetes, hypertension, Hyperlipidemia, NSAID intake, multivitamin use, Hypolipidemic drug intake and intake of mineral supplements; **Abbreviations:** IMD, index of multiple deprivation; BMI, body mass index; NSAID, nonsteroidal anti-inflammatory drug.

Stratified by drug class, the effect of modification by diabetes and hyperlipidemia were evaluated. There was a significant interaction with diabetes among users of anticoagulants (P for interaction = 0.019). Given that warfarin was the predominant anticoagulant (other anticoagulants were not analyzed due to insufficient sample size), a subgroup analysis focusing on warfarin confirmed that this interaction remained statistically significant (P for interaction = 0.015). Specifically, for participants without diabetes, conventional use of warfarin was associated with a 64% increased risk of developing non-biliary AP (HR, 1.64; 95% CI, 1.15-2.35), whereas there was no statistically significant association for participants with diabetes. Furthermore, hyperlipidemia did not have a significant modifying effect on anticoagulants. For users of antiplatelet agents, including low-dose aspirin, clopidogrel, and dipyridamole, there was no significant interaction with diabetes or hyperlipidemia (Supplementary Table 3).

### Sensitivity analysis

The association between the use of anticoagulants and the risk of developing nonbiliary AP remained stable after further adjustment for a history of cardiovascular disease (HR, 1.31; 95% CI, 1.08-1.61). After excluding participants with nonbiliary AP that developed within the first year of follow-up, the association remained statistically significant (HR, 1.25; 95% CI, 1.01-1.54; Supplementary Table 4). In a sensitivity analysis using a propensity score-matched cohort of 55,694 participants (Supplementary Table 5), regular use of antithrombotic drugs was associated with a 37% increased risk of developing nonbiliary AP (HR, 1.37; 95% CI, 1.01-1.86).

## DISCUSSION

In this prospective cohort study, using data from >430,000 participants, regular use of antithrombotic drugs was associated with a 31% increased risk of developing nonbiliary AP, after adjusting for potential confounding factors. In-depth analysis revealed significant heterogeneity in the level of risk; the increased risk was primarily concentrated in users of clopidogrel (HR, 1.53), whereas warfarin had only a trend towards increased risk, and aspirin and dipyridamole did not have a significant risk. Notably, the association remained robust in a sensitivity analysis, with further adjustment for a history of cardiovascular diseases. This consistency strengthens the argument for there being an independent association between the use of antithrombotic drugs and the risk of developing nonbiliary AP, beyond the influence of underlying cardiovascular conditions.

To the best of our knowledge, this is the first prospective cohort study, in which the association between the use of antithrombotic drugs and the risk of developing nonbiliary AP has been investigated. Although previous studies, on the association between the use of antithrombotic drugs and disorders of the stomach, small intestine [15,16], and liver [17,18], and their potential adverse effects on the pancreas, have received limited attention. In 2015, based on a population-based case-control study in Taiwan, Lai et al. reported a significantly increased risk of developing AP in patients actively using clopidogrel (adjusted odds ratio, 8.46; 95% CI, 5.25–13.7). The study was a population-based case-control design and included 5,644 patients with AP, significantly enhancing the reliability of the results [13]. The results strongly support the findings of the present study. Moreover, in the present study, diabetes mellitus significantly modified the association between warfarin and the risk of developing nonbiliary AP, and these findings provide important evidence for personalized risk assessment in clinical practice.

From the results of the present study, a potential mechanism for the increased risk of developing nonbiliary AP in users of clopidogrel, is proposed. Clopidogrel, as a P2Y12 receptor antagonist, might contribute to pancreatic injury by affecting P2Y12 receptors on pericytes in the pancreatic microcirculation. Recent studies have shown that P2Y12 receptors are also expressed on vascular smooth muscle cells (VSMCs), and their activation inhibits the cAMP/protein kinase A pathway [19]. This inhibition effect leads to dephosphorylation of the cytoskeletal regulator, cofilin, which promotes actin depolymerization and enhances cell motility [20,21]. Given the high degree of homology between VSMCs and pericytes, and the established expression of P2Y12 receptors in pericytes [22], pancreatic pericytes also likely to express functional P2Y12 receptors. Signals transmitted through these receptors regulate cytoskeletal proteins such as actin-depolymerizing factors, thereby maintaining pericyte adhesion capacity and cytoskeletal plasticity. Clopidogrel antagonism disrupts this regulation, reducing cytoskeletal remodeling and increasing pericyte rigidity. This disruption effect compromises their capillary support function and connections with endothelial cells, thereby impairing the integrity of the microvascular barrier. The consequent increase in vascular permeability promotes the extravasation of inflammatory mediators and leukocyte infiltration, ultimately triggering or exacerbating local pancreatic inflammation [23,24]. In contrast, aspirin inhibits thromboxane A2 synthesis by acetylating platelet COX-1, an action that is highly restricted to platelets [25]. Dipyridamole enhances the intracellular cAMP level by inhibiting phosphodiesterase, thereby suppressing platelet activation and promoting vasodilation, and it potentiates these effects by elevating the adenosine level [26]. As neither drug directly interferes with the P2Y12 receptor signaling pathway, there were no significant associations with the risk of developing nonbiliary AP.

Subgroup analyses revealed that the association between the use of warfarin and the risk of developing nonbiliary AP was modified by the presence of type 2 diabetes. This association was statistically significant in individuals without diabetes, and it was not significant in individuals with diabetes. The mechanism underlying the effect modification remains unclear, and it can potentially be attributed to the balance between vascular injury and anticoagulation. As a vitamin K antagonist, warfarin inhibits the synthesis of multiple vitamin K-dependent proteins, including matrix Gla protein (MGP). MGP is a key factor in maintaining vascular homeostasis and suppressing calcification [27,28]. Warfarin may impair pancreatic microvascular function by inducing vascular calcification through inhibition of MGP, ultimately increasing the susceptibility of pancreatic tissue to inflammation. However, diabetes status often co-exists with a hypercoagulable state, which itself constitutes a risk factor for pancreatic microcirculatory dysfunction and AP [29–31]. In this context, the anticoagulant effect of warfarin may partially improve pancreatic microcirculation, thereby offsetting its potential risks via the MGP pathway. Conversely, the direct injury mechanism of warfarin predominates in individuals without diabetes and normal coagulation function. Furthermore, the extremely high baseline risk of pancreatitis in patients with diabetes may partially mask the additional relative risk posed by warfarin [32]. Further studies are required to explore the underlying mechanisms.

This study had several strengths, including its prospective design, large sample size, and extended follow-up period. Regular use of antithrombotic drugs is associated with a higher risk of developing nonbiliary AP, after controlling for several potential confounding factors, including lifestyle factors, use of medication, and health conditions related to the use of antithrombotic drugs. The scale and depth of the dataset permitted rigorous adjustment of potential confounders.

However, this study had several limitations. First, in the UK Biobank, data on specific details, such as formulation, dosage, frequency, and duration of paracetamol administration were not documented. This limitation hinders further analysis, and it may introduce potential bias. Second, information on the use of antithrombotic drugs was collected only once, at baseline, which prevented an assessment of how changes in exposure over time might affect the risk of developing liver cancer. Third, information on the severity of AP (according to the Revised Atlanta Classification) was not available in the databases. This precluded further stratified analyses based on severity of disease. Finally, as an observational study, a causal relationship cannot be definitively established, and residual confounding from unmeasured variables cannot be ruled out. These inherent limitations highlight the requirement for further epidemiological and mechanistic studies.

## CONCLUSIONS

This large prospective cohort study has identified that regular use of antithrombotic drugs, particularly clopidogrel, is associated with an increased risk of developing nonbiliary AP. Furthermore, underlying metabolic status was identified as a key modifier of this risk. Notably, diabetes status had a negative modifying effect on warfarin-associated risk, which was more pronounced in individuals without diabetes. This counter-intuitive finding underscores the necessity for personalized risk assessment in clinical decision-making for antithrombotic therapy, with vigilance warranted in seemingly “healthy” cohorts. Future studies are required to elucidate the underlying biological mechanisms, and to validate these findings at the level of specific drug subtypes in high-quality cohorts.

## LIST OF ABBREVIATIONS

AP: Acute Pancreatitis
BMI: Body Mass Index
CI: Confidence Interval
HR: Hazard Ratio
ICD-10: International Classification of Diseases-10
IMD: Index of Multiple Deprivation
Met: Metabolic Equivalent of Task
MGP: Matrix Gla Protein
NSAID: Nonsteroidal Anti-Inflammatory Drug
PSM: Propensity Score Matching
SD: Standard Deviation
UKB: UK Biobank
VSMCs: vascular smooth muscle cells

## DECLARATIONS

### Ethics approval and consent to participate

The UK Biobank has received ethical approval from the North West Multi-centre Research Ethics Committee, the England and Wales Patient Information Advisory Group and the Scottish Community Health Index Advisory Group. All participants provided written informed consent prior to enrolment, and the analysis used anonymous data.

### Consent for publication

Not Applicable.

### Availability of data and materials

All data used in this study was obtained from publicly available datasets UK biobank (UKB). To access the data used in this study, researchers can apply for access to the UKB through their website (https://www.ukbiobank.ac.uk/enable-your-research/apply-for-access).

### Clinical Trial Number

Not Applicable.

### Competing interests

The authors have no conflicts of interest to declare.

### Funding

This study was supported by the Major Program of the Gansu Province Joint Scientific Research Fund (23JRRA1488); Major Scientific and Technological Innovation Project of Gansu Provincial Health Industry (GSWSQNPY2024-10); and the Lanzhou Talent Innovation and Entrepreneurship Project (2022-3-42).

### Author Contributions

Wei CY and Mi NN contributed to conceptualization and design; Wei CY, Mi NN, Zhao JY, Li PF, An ZP and Chen S contributed to material preparation, data acquisition; Yuan JQ, Lin YY and Yue P analyzed the data. Meng WB: supervised our study. all authors participated in the writing and final approval of the manuscript.

## Acknowledgements

The authors thank the participants and staff of the UKB for their dedication and contribution to the research.

## Notes

### Competing Interest Statement

The authors have declared no competing interest.

### Summary of Updates

Corrections have been made to linguistic errors and some unclear phrasing.

