## supplementary material for "Association between regular antithrombotic drugs use and Nonbiliary Acute Pancreatitis risk: a prospective cohort study"

**Correspondence**

**Supplementary Table 1.** Assessment of main exposures.

**Supplementary Table 2.** Covariates about Physical activity and Dietary characteristics.

**Supplementary Table 3.** Effect modification by diabetes and hyperlipidemia on the **Supplementary** association between antithrombotic drug use and Nonbiliary Acute **Pancreatitis** risk.

**Supplementary Table 4.** Sensitivity analysis regarding risk of Nonbiliary Acute Pancreatitis according to Antithrombotic use.

**Supplementary Table 5.** Baseline characteristics according to Antithrombotic use after 1:4 propensity score matching.

**Supplementary Figure 1.** Flowchart of participant selection.

**Supplementary Table 1.** Assessment of main exposures.

| **Description** | **Questionnaire** | **Responses** | **Field ID** |
| --- | --- | --- | --- |
| Prescription medications taking | Do you regularly take any other PRESCRIPTION medications? (Do not forget medications such as puffers or patches) | SELECT one of 4 from  1 : Yes - you will be asked about this later by an interviewer  0 : No  1 : Do not know -3 : Prefer not to answer | 2492 |

| **Field ID** | **Meaning** | **Data-Coding 4** |
| --- | --- | --- |
| 20003 | Warfarin | 1140888266 |
|  | Sodium Warfarin | 1140910832 |
|  | Marevan 0.5mg tablet | 1141164760 |
|  | Dipyridamole | 1140861778 |
|  | Persantin 25mg tablet | 1140861780 |
|  | Angettes 75mg tablet | 1140861804 |
|  | Aspirin 75mg tablet | 1140861806 |
|  | Disprin CV 100mg m/r tablet | 1140861808 |
|  | Clopidogrel | 1141168318 |
|  | Plavix 75mg tablet | 1141168322 |

**Supplementary Table 2. Covariates about Physical activity and Dietary characteristics.**

| **Healthy diet items** | **Questionnaire** | **Healthy diet** | **Unhealthy diet** |
| --- | --- | --- | --- |
| Fruit | “About how many of …. would you eat per day?” Separate questions for pieces of fresh and dried fruit. | ≥ 4.5 pieces/day | < 4.5 pieces/day |
| Vegetable intake | “About how many of …. would you eat per day?” Separate questions for tablespoons of salad or cooked/raw vegetables. | ≥4.5 servings/day | < 4.5 servings/day |
| Total fish intake | “How often do you eat oily fish? (e.g., sardines, salmon, mackerel, herring)” | ≥2 times/week | < 2 times/week |
| Red meat intake | “How often do you eat…?” Separate questions for Beef/lamb or mutton/pork (excluding processed meats such as ham or bacon). | ≤5 times/week | >5 times/week |
| Processed meat intake | “How often do you eat processed meats (such as bacon, ham, sausages, meat pies, kebabs, burgers, chicken nuggets)?” | ≤2 times/week | >2 times/week |

Note: Physical activity was measured using the validated short-form International Physical Activity Questionnaire[1].

Dietary characteristics (From touchscreen questionnaire at baseline. Healthy diet patterns were adapted from the American Heart Association Guidelines and were defined as meeting 2 or more healthy diet items).

Tablespoons of vegetables were considered one serving. The healthy diet score was dichotomized as 1 = at least two healthy food items, 0 = fewer than two healthy food items.

**Supplementary Table 3. Effect modification by diabetes and hyperlipidemia on the association between antithrombotic drug use and Nonbiliary Acute Pancreatitis risk.**

| **Drug** **Class** | **Subgroup** | **HR (95%CI)** | **P Value** | **P for** **Interaction** |
| --- | --- | --- | --- | --- |
| **Antithrombotic** |  |  |  |  |
|  | Diabetes: Yes | 1.08 (0.78-1.50) | 0.644 | 0.01 |
|  | Diabetes:No | 1.52 (1.19-1.96) | 0.001 |  |
|  | Hyperlipidemia: Yes | 1.23 (0.99-1.53) | 0.063 | 0.044 |
|  | Hyperlipidemia: No | 1.96 (1.22-3.15) | 0.006 |  |
| **Anticoagulants** |  |  |  |  |
|  | Diabetes: Yes | 0.97 (0.54-1.72) | 0.905 | 0.019 |
|  | Diabetes:No | 1.68 (1.18-2.39) | 0.004 |  |
|  | Hyperlipidemia: Yes | 1.26 (0.89-1.79) | 0.196 | 0.117 |
|  | Hyperlipidemia: No | 1.98 (1.09-3.61) | 0.026 |  |
| **Warfarin** |  |  |  |  |
|  | Diabetes: Yes | 0.89 (0.49-1.63) | 0.716 | 0.015 |
|  | Diabetes: No | 1.64 (1.15-2.35) | 0.007 |  |
|  | Hyperlipidemia: Yes | 1.23 (0.87-1.76) | 0.247 | 0.179 |
|  | Hyperlipidemia: No | 1.82 (0.97-3.42) | 0.062 |  |
| **Antiplatelets** |  |  |  |  |
|  | Diabetes: Yes | 1.13 (0.76-1.68) | 0.549 | 0.099 |
|  | Diabetes: No | 1.40 (1.00-1.96) | 0.05 |  |
|  | Hyperlipidemia: Yes | 1.21 (0.92-1.59) | 0.164 | 0.218 |
|  | Hyperlipidemia: No | 1.89 (0.88-4.02) | 0.1 |  |
| **Clopidogrel** |  |  |  |  |
|  | Diabetes: Yes | 1.38 (0.82-2.32) | 0.228 | 0.214 |
|  | Diabetes: No | 1.69 (1.06-2.68) | 0.027 |  |
|  | Hyperlipidemia: Yes | 1.49 (1.04-2.13) | 0.028 | 0.522 |
|  | Hyperlipidemia: No | 2.10 (0.52-8.56) | 0.298 |  |

Note: The model was adjusted for age, gender, ethnicity, the Index of Multiple Deprivation (IMD), BMI, smoking status, alcohol intake, physical activity, dietary characteristics, type 2 diabetes, hypertension, hyperlipidemia, NSAID intake, hypolipidemic drug intake, multivitamin use, and mineral supplement intake. Formal interaction analysis for the subgroup of patients using DOACs or LMWH was not performed due to an insufficient sample size.

**Supplementary Table 4. Sensitivity analysis regarding risk of Acute pancreatitis according to Antithrombotic use.**

| **Sensitivity analysis** | **No. of Acute pancreatitis** | **No. of participants** | **Adjusted HR (95%CI)** | ***p*-value** |
| --- | --- | --- | --- | --- |
| **1.** Added overall History of cardiovascular and cerebrovascular disease to model 3 as model 4 for additional adjustment. | | | | |
| Non-Antithrombotic use | 2080 | 420614 | 1.00 (Reference) |  |
| Antithrombotic use | 109 | 11140 | 1.31(1.08-1.61) | 0.007 |
| **2.** Excluding cholelithiasis participants diagnosed within 1 years after baseline (**N=428,986**). | | | | |
| Non-Antithrombotic use | 1997 | 418028 | 1.00 (Reference) |  |
| Antithrombotic use | 98 | 10958 | 1.21 (1.16-1.26) | <0.001 |
| **3.** Propensity score matching cohort between Antithrombotic use and non-Antithrombotic use (**1:4 matching, N=55249**). | | | | |
| Non-Antithrombotic use | 336 | 44218 | 1.00 (Reference) |  |
| Antithrombotic use | 109 | 11031 | 1.36 (1.10-1.69) | 0.005 |

Note: Except for sensitivity analysis 1, which further adjusted for History of cardiovascular and cerebrovascular disease, all other adjusted HRs were adjusted for the following covariates: age, gender, ethnicity, IMD, BMI, smoking status, alcohol intake, physical activity, dietary characteristics, type 2 diabetes, hypertension, hyperlipidemia, NSAID intake, Hypolipidemic drug intake,multivitamin use and intake of mineral supplements. HR, Hazard ratio; CI, confidence interval; IMD, index of multiple deprivation; BMI, body mass index; NSAID, nonsteroidal anti-inflammatory drug.

**Supplementary Table 5. Baseline characteristics according to Antithrombotic use after 1:4 propensity score matching.**

|  | **Overall** | **Antithrombotic use** | |
| --- | --- | --- | --- |
|  | N = 55694 | N = 44554 | N = 11140 |
| **Age (mean (SD))** | 61.77 (6.11) | 61.80 (6.07) | 61.67 (6.26) |
| **Male (%)** | 38911 (69.9) | 31112 (69.8) | 7799 (70.0) |
| **White race(%)** | 52499 (94.3) | 41997 (94.3) | 10502 (94.3) |
| **IMD (mean (SD))** | 19.31 (15.12) | 19.29 (15.07) | 19.40 (15.33) |
| **BMI (mean (SD))** | 29.22 (5.04) | 29.22 (5.01) | 29.21 (5.18) |
| **Smoking (%)** | 32190 (57.8) | 25736 (57.8) | 6454 (57.9) |
| **Drinking (%)** |  |  |  |
| Never | 13934 (25.0) | 11124 (25.0) | 2810 (25.2) |
| Low | 5505 ( 9.9) | 4401 ( 9.9) | 1104 ( 9.9) |
| Moderate | 23770 (42.7) | 19019 (42.7) | 4751 (42.6) |
| High | 12423 (22.3) | 9962 (22.4) | 2461 (22.1) |
| **MET_g (%)** |  |  |  |
| high | 15259 (27.4) | 12212 (27.4) | 3047 (27.4) |
| low | 9491 (17.0) | 7562 (17.0) | 1929 (17.3) |
| moderate | 16997 (30.5) | 13618 (30.6) | 3379 (30.3) |
| **HealthyDiet (%)** | 28768 (51.7) | 23000 (51.6) | 5768 (51.8) |
| **NASIDS (%)** | 3615 ( 6.5) | 2916 ( 6.5) | 699 ( 6.3) |
| **Hypolipidemic drug (%)** | 40672 (73.0) | 32530 (73.0) | 8142 (73.1) |
| **Vitamin (%)** | 8130 (14.6) | 6494 (14.6) | 1636 (14.7) |
| **Mineral (%)** | 8005 (14.4) | 6409 (14.4) | 1596 (14.3) |
| **Diabetes (%)** | 14783 (26.5) | 11755 (26.4) | 3028 (27.2) |
| **Hypertension (%)** | 49330 (88.6) | 39511 (88.7) | 9819 (88.1) |
| **Hyperlipidemia (%)** | 47328 (85.0) | 37965 (85.2) | 9363 (84.0) |
| **Myocardial Infarction (%)** | 2239 ( 4.0) | 1800 ( 4.0) | 439 ( 3.9) |
| **Coronary Heart Disease (%)** | 3066 ( 5.5) | 2467 ( 5.5) | 599 ( 5.4) |
| **PVD (%)** | 656 ( 1.2) | 520 ( 1.2) | 136 ( 1.2) |
| **AF (%)** | 937 ( 1.7) | 749 ( 1.7) | 188 ( 1.7) |
| **stroke (%)** | 932 ( 1.7) | 745 ( 1.7) | 187 ( 1.7) |

Note: Abbreviations: SD, standard deviation; NSAID, nonsteroidal anti-inflammatory drug; PVD: myocardial infarction, AF: peripheral vascular disease.

Values are numbers (percentages) unless stated otherwise.

**Supplementary Figure 1. Flowchart of participant selection.**

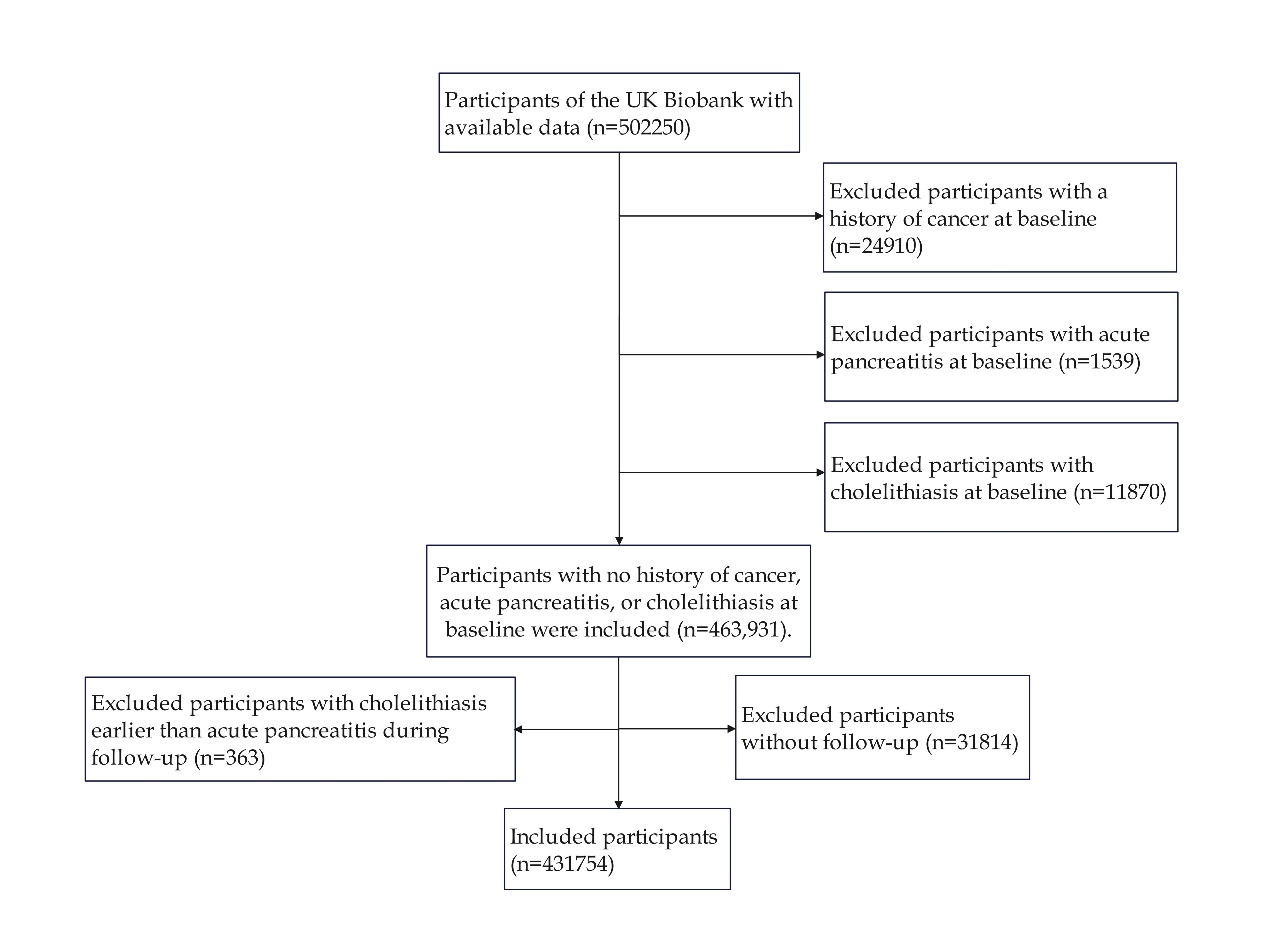
